# Comparison and aggregation of event sequences across ten cohorts to describe the consensus biomarker evolution in Alzheimer’s disease

**DOI:** 10.1101/2021.11.14.21266316

**Authors:** Sepehr Golriz Khatami, Yasamin Salimi, Martin Hofmann-Apitius, Neil P. Oxtoby, Colin Birkenbihl, for the Alzheimer’s Disease Neuroimaging Initiative, the Japanese Alzheimer’s Disease Neuroimaging Initiative, the Alzheimer’s Disease Repository Without Borders Investigators

## Abstract

**Background:** Previous models of Alzheimer’s disease (AD) progression were primarily hypothetical or based on data originating from single cohort studies. However, cohort datasets are subject to specific inclusion and exclusion criteria that influence the signals observed in their collected data. Furthermore, each study measures only a subset of AD relevant variables. To gain a comprehensive understanding of AD progression, the heterogeneity and robustness of estimated progression patterns must be understood, and complementary information contained in cohort datasets be leveraged.

**Methods:** We compared ten event-based models that we fit to ten independent AD cohort datasets. Additionally, we designed and applied a novel rank aggregation algorithm that combines partially overlapping, individual event sequences into a meta-sequence containing the complementary information from each cohort.

**Results:** We observed overall consistency across the ten event-based model sequences (Kendall’s tau correlation coefficient of 0.78±0.13), despite variance in the positioning of mainly imaging variables. The changes described in the aggregated meta-sequence are broadly consistent with current understanding of AD progression, starting with cerebrospinal fluid amyloid beta, followed by memory impairment, tauopathy, FDG-PET, and ultimately brain deterioration and impairment of visual memory.

**Conclusion:** Overall, the event-based models demonstrated similar and robust disease cascades across independent AD cohorts. Aggregation of data-driven results can combine complementary strengths and information of patient-level datasets. Accordingly, the derived meta-sequence draws a more complete picture of AD pathology compared to models relying on single cohorts.

## 1. Background

Alzheimer’s disease, in combination with its clinical manifestation/syndrome (AD) **[1]**, is a progressive, multifaceted disease whose cognitive symptoms surface years after disease onset **[2]**. In order to identify crucial opportunities for medical interventions that could potentially prevent or delay symptoms, it is vital to understand the temporal relationship of pathological changes underlying the progressive nature of AD. To this end, cognitive assessments and a wide range of biomarkers, including cerebrospinal fluid (CSF) markers and neuroimaging derived measures, have been established to monitor the disease’s progression. Measuring these markers enables the observation of biochemical, structural, functional, and cognitive changes that occur as the disease progresses **[3]** and the resulting data can build the basis for data-driven approaches that aim to determine the relative temporal dependencies between biomarkers and cognitive symptoms **[4]**. Previously, a variety of data-driven models have been developed with the aim of accomplishing this task **[5-10]**.

One model archetype that has found wide success in the context of neurodegenerative diseases **[11-14]** and AD specifically **[15]** is the event-based model (EBM) **[13]**. It is a data-driven probabilistic generative model that characterises the progression of a disease in the form of a single sequence of events which describes the relative order of measured markers turning from a normal state to a diseased state (i.e., abnormal state). Such event sequences carry the benefit that they are highly interpretable and, although describing disease progression, can already be learned from cross-sectional cohort study data. Previously, EBMs have been used to derive event sequences **[13]**, stage subjects in their disease progression **[15]**, predict conversion from one clinical stage to the other (i.e., CU to mild cognitive impairment (MCI), or MCI to AD) **[16]**, and uncover disease phenotypes with distinct temporal progression patterns.

To build an EBM, patient-level data are needed on which the model can be fit. In recent decades, an increasing number of observational cohort studies have released their collected data for research purposes, including the Alzheimer’s Disease Neuroimaging Initiative (ADNI) **[17]**, the European Prevention of Alzheimer’s Dementia (EPAD) **[18]**, and AddNeuroMed **[4]**. So far, however, only a few studies in the AD domain have applied EBMs to data from other cohorts besides ADNI **[19,20]**. Previous work evaluating data-driven progression modeling based on cohort datasets has shown that the participant recruitment procedures can introduce cohort-specific systematic statistical biases into the collected data **[21]**, which, in turn, can bias the estimation of disease progression **[22]**. Therefore, it is necessary to replicate and validate data-driven results in independent cohorts to ensure robust conclusions. Consequently, it remains unclear whether event sequences determined from one cohort dataset would generalize beyond the discovery cohort itself and, further, if sequences generated across several cohorts were concordant among each other. Simultaneously, gaining a comprehensive event sequence combining all relevant AD biomarkers, cognitive assessments, and functional scores is infeasible, since cohort studies can only measure a limited set of variables that are often only partially overlapping between them **[23]**. In theory, however, this allows for an estimation of individual event sequences from distinct cohorts which cover complementary sets of markers. Aggregating results across cohorts would harness this complementary information by assembling a meta-sequence that provides a more complete picture of the development and progression of AD.

In this work, we present a systematic, in-depth comparison of AD event sequences derived from ten independent landmark cohort studies to investigate the generalizability and robustness of EBM derived AD progression patterns. Furthermore, we designed a novel rank aggregation algorithm which we used to aggregate the event sequences into a single meta-sequence, thereby fusing the complementary information in all variables assessed across the studies. Our work harnesses the heterogeneity in cohort study designs and measurements to produce a meta-sequence providing a more complete, and robust, picture of the temporal order of pathological marker changes in AD progression.

## 2. Methods

### 2.1. Investigated cohort datasets

We selected ten independent AD cohort studies for our analysis by systematically exploring suitable datasets using the ADataViewer **[23]**. The prerequisite for including a cohort into our analysis was that 1) diagnostic staging into CU, MCI, and AD was performed **[24]**, 2) sufficient cross-sectional data was available for all diagnostic stages, and 3) multiple data modalities were collected. The cohorts that were ultimately selected are presented in **Table 1**.

**Table 1.** Selected cohorts, their number of participants per disease stage, and their number of considered variables.

| Cohort | Consortium | CU | MCI | AD | Total | Number of biomarkers |
| --- | --- | --- | --- | --- | --- | --- |
| ADNI [17] | The Alzheimer's Disease Neuroimaging Initiative | 30 | 69 | 36 | 135 | 18 |
| JADNI [25] | Japanese Alzheimer's Disease Neuroimaging Initiative | 17 | 30 | 67 | 114 | 18 |
| AIBL [26] | The Australian Imaging, Biomarker Lifestyle Flagship Study of Ageing | 92 | 23 | 13 | 128 | 10 |
| NACC [27] | The National Alzheimer's Coordinating Center | 24 | 21 | 45 | 90 | 16 |
| ANM [28] | AddNeuroMed | 123 | 135 | 145 | 403 | 7 |
| EMIF-1000 [29] | European Medical Information Framework | 47 | 229 | 53 | 329 | 9 |
| EDSD [30] | European DTI Study on Dementia | 38 | 22 | 32 | 92 | 12 |
| ARWIBO [31] | Alzheimer's Disease Repository Without Borders | 226 | 97 | 44 | 367 | 10 |
| OASIS-1 [32],<br>OASIS-2 [33] | Open Access Series of Imaging Studies | 135 | 70 | 30 | 235 | 7 |
| WMHAD [34] | White Matter Hyperintensities in Alzheimer's Disease | 19 | 27 | 42 | 88 | 13 |

### 2.2 Variable selection

We aimed at including a wide spectrum of variables to uncover the temporal relationship across multimodal markers of AD pathology that capture, for example, different biochemical, cognitive, or structural changes. In order to include a specific variable, it must have been measured in all diagnostic groups of the respective study. Further, only a minimal amount of missing values was tolerable, as participants with missing values in any of the ultimately selected variables had to be excluded from the analysis. This led to a trade-off between the inclusion of an increasing number of variables and the amount of participants available for analysis. In total, 36 unique variables were selected from different data modalities covering neuropsychological and cognitive tests, CSF markers, and MRI derived brain region volumes. The complete list of selected biomarkers and their corresponding modality are presented in **Table 2**. The number of variables per cohort is given in **Table 1**.

**Table 2.** The selected biomarkers and their corresponding abbreviations.

| Modality | Biomarker | Abbreviation | Cohorts containing variable |
| --- | --- | --- | --- |
| Clinical assessments | Clinical Dementia Rating Sum of Boxes | CDRSB | 3 |
|  | Neuropsychiatric Inventory | NPI | 2 |
|  | Logical Memory - Delayed Recall Total Number of Story Units Recalled | LDEL | 5 |
|  | Alzheimer's Disease Assessment Scale (13-items) | ADAS13 | 2 |
|  | Alzheimer's Disease Assessment Scale (11-items) | ADAS11 | 2 |
|  | Mini-Mental State Examination | MMSE | 10 |
|  | Logical Memory - Immediate Recall Total Number of Story Units Recalled | LIMM | 6 |
|  | Trail Making Test-B | TRABS | 2 |
|  | Digit-Symbol Coding Test | DIGITS | 2 |
|  | California Verbal Learning Test Delayed Raw Score | LIDE | 1 |
|  | Category Fluency (animals - fruits/vegetables) | CATFLU | 3 |
|  | Figure Copy | FIGC | 3 |
|  | California Verbal Learning Test Recall Raw Score | LIRE | 2 |
|  | Figure recall | FIGR | 2 |
|  | C/D Stroop Test Raw | STROOP | 1 |
|  | Short Term Memory | STM | 1 |
|  | Language | LANGU | 1 |
|  | Perceptual Orientation | ORIENT | 2 |
|  | Mental Manipulation | MENMA | 1 |
|  | Attention | ATTEN | 1 |
|  | Clock Drawing Test Total Score | CLKS | 2 |
|  | Executive Memory | EXECUTIVE | 1 |
|  | Word List Learning Trial | LICOR | 1 |
|  | Boston Naming Test Score | BNTS | 2 |
|  | Digit Symbol Substitution Test | WAIS | 2 |
| CSF markers | Amyloid-β | ABETA | 4 |
|  | Total Tau | TAU | 4 |
|  | Phosphorylated Tau (p-Tau) | PTAU | 4 |
| Imaging markers | Entorhinal Volume | ENTOR | 8 |
|  | Hippocampal Volume | HIPPO | 8 |
|  | Fusiform Volume | FUSIF | 8 |
|  | Ventricles Volume | VENT | 8 |
|  | MiddleTemporal Volume | MIDTEPM | 8 |
|  | Accumulated CSF in the Brain | CSFVOL | 5 |
|  | Fluorodeoxyglucose Positron Emission Tomography (FDG PET) | FDG | 2 |

### 2.3. Participants

An available diagnosis of a participant as either CU, MCI, or AD was prerequisite for inclusion. Further, any participant with a diagnosis of cognitive impairment that was not linked to AD by the respective study’s clinicians was excluded. Further, only participants with complete data across all selected biomarkers could be used in our modelling approach. The number of participants per cohort and diagnostic group are described in **Table 1**.

### 2.4. Progression modeling via event-based models

The EBM derives a probabilistic sequence from patient-level data that describes the temporal order in which measured values of variables turn from a normal to an abnormal state. Each of these transitions is called an event. In this context, normality or abnormality are defined non-parametrically using kernel density estimation mixture modelling on the empirical values of the modeled cohort’s CU and AD populations, respectively **[35]**. This probabilistic allocation of measurements into two groups, allows study participants (in particular, patients) to have a mix of occurred and non-occurred events across all measurements which lays the foundation to estimate the most likely event sequence. Here, the EBM assumes that the biomarkers monotonically change towards abnormality as the disease progresses and that this process is irreversible. Furthermore, there are no a priori assumptions regarding predefined disease stages, cut points determining the abnormality of biomarkers, or the temporal relationship between them. The most likely sequence of events *S* is then estimated by maximizing the likelihood *pr*(*X*|*S*) **(Equation 1)**, where variable measurements are denoted by *x* ∈ *X* for *i* ∈ *M* markers and *j* ∈ *N* indicates the individual samples.

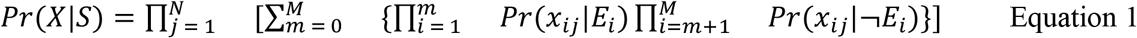

Here Pr (*x*_*ij*_ |*Ei*)and Pr(*x*_*ij*_ |¬*E*_*i*_) describe the probability of observing the value of *x* given that the event *E*_*i*_ (i.e., variable *x* turning abnormal) has, or has not, occurred, respectively. For more details, we refer to the Supplementary Material and the original publication of the KDE EBM by Firth et al. **[35]**. The derived mixture models per cohort and measurement are presented in **Figure S3**.

To quantify the similarity of distinct event sequences, we calculated the pairwise Kendall’s tau rank correlation coefficient (KTC) across complete sequences and the Bhattacharrya coefficient (*BC*) for specific events as explained in Oxtoby et al. **[12]**. An average KTC that is close to 1 and shows low standard deviation across the cohorts would indicate high concordance. An average BC close to 1 implies high similarity in the positional variance of ranks while the BC amounts to 0 for completely different patterns.

### 2.5. Meta-sequence generation

To generate a meta-sequence, we propose a method that combines individual event sequences (called base-sequences) stemming from independent datasets. We assemble a meta-sequence in a two step procedure: first, building on the ideas presented in **[36]** and **[37]**, we generate all possible sequences comprising *k* variables that are randomly drawn from the union of variables encountered in the base-sequences (with *k* < total number of variables). The generated sequence with the minimum average distance to all base-sequences is selected as a starting point for the next step. In step two, this starting sequence is extended by iteratively adding more variables to it, such that the average distance between the altered sequence and all base-sequences remains minimal. Here, the new variable is not necessarily added to the end of the sequence but all possible positions are considered. This process is repeated until all variables have been included into the sequence which finally forms the aggregated meta-sequence. Splitting the algorithm in two steps (an exhaustive search for the first *k* variables followed by the greedy insertions) was necessary, as the search space (i.e., all possible meta-sequences) grows exponentially with the number of variables in the base-sequences. Further explanations about the algorithm, the handling of partially overlapping lists, and access to the corresponding python code are provided in the Supplementary Material and **Figure S1**.

For this work, we generated a meta-sequence considering only variables which were present in at least three cohorts (**Table 2**) and set *k* equal to eight. The distance metric chosen was Spearman’s footrule distance which takes the absolute difference in positions of variables into account.

## 3. Results

### 3.1. Comparing event sequences derived from multiple cohort studies

We observed good consistency with respect to the position of events across all cohorts’ sequences which resulted in an average KTC of 0.78 ± 0.13 (pairwise Kendall’s tau rank correlations are presented in **Table S3**; sequence similarity is also indicated visually through an approximately diagonal line of the event ranks from top-left to bottom-right in **Figure 1**). The exception to this was the WMHAD cohort, for which a substantial deviation from the other sequences was detected. In most cohorts’ sequences, cognitive assessment scores ranked highly, before CSF markers, which were followed by MRI-derived brain volumes in the lower ranks. Only for NACC and JADNI, a non-cognitive marker, namely beta amyloid (ABETA), was determined to be the first event.

**Figure 1.**
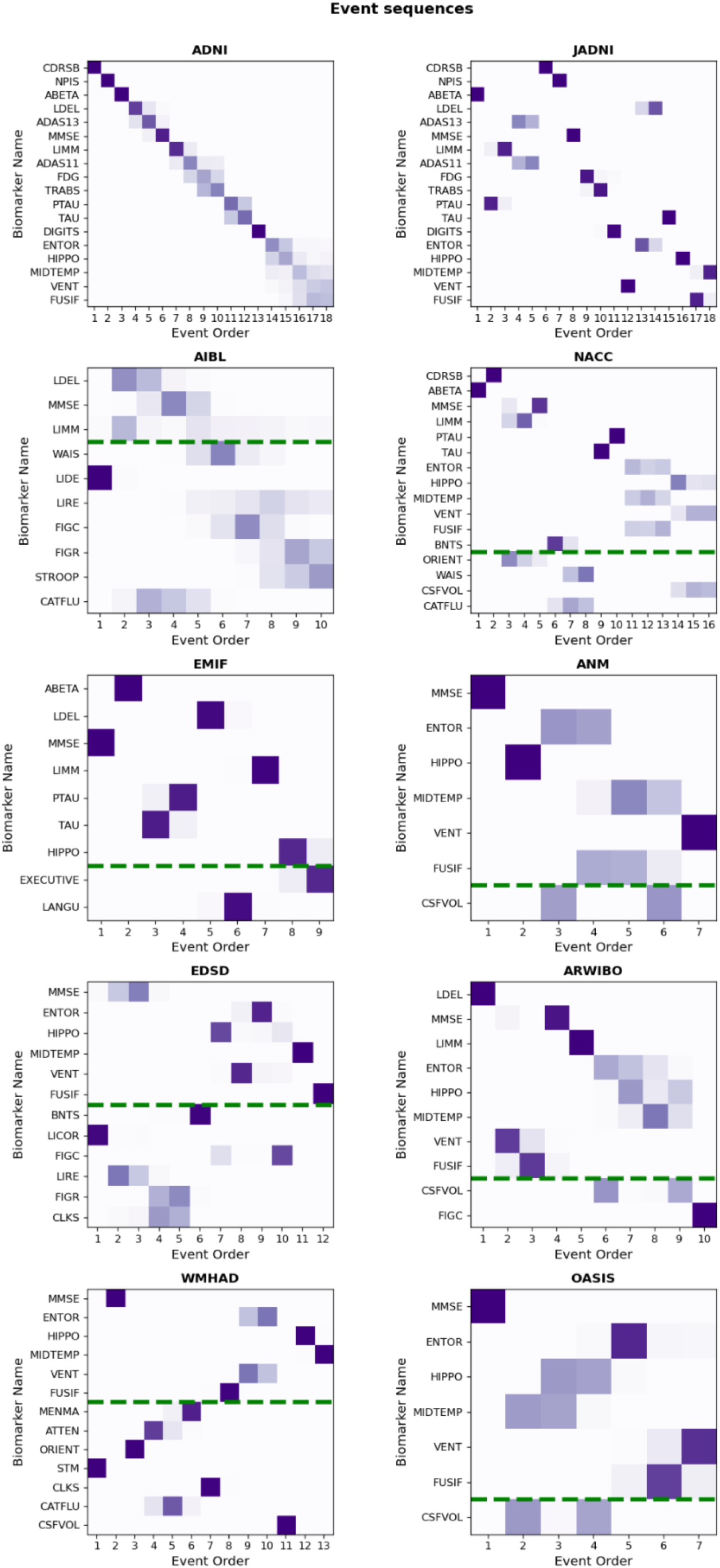
Individual event sequences estimated from the ten investigated cohorts. For comparison of relative event positions, the y-axes follow the ADNI sequence. Common events between ADNI and the other cohorts are presented above a dashed green line. The closer the sequences are to ADNI, the more diagonal the probabilistic position (colored squares) will align from top-left to bottom-right. The event sequences in their original form are presented in **Figure S2**.

The relative order among clinical assessments measuring different cognitive domains (e.g., memory, language, visuospatial, executive) was consistent across all cohorts (**Table S1**). The cognitive impairment in all investigated cohorts started with memory dysfunction detected by logical memory tests (e.g., LDEL and LIMM), proceeded with language impairments exposed by tests such as the BNT and CATFLU, visual dysfunction identified through the CLKS or FIGC, and finally executive dysfunction recognized by, for example, the DIGIT and WAIS.

Amongst the cohorts where CSF biomarkers had been measured (ADNI, JADNI, EMIF, NACC), the relative positions of these biomarkers, in particular of tau (TAU) and phosphorylated tau (PTAU), varied. While ABETA consistently placed among the top three positions in all of these cohorts’ sequences, TAU and PTAU were either found in early positions as well (JADNI and EMIF), or in the middle of the sequence (ADNI and NACC). However, in all cases PTAU and TAU were direct neighbors, indicating the consistent, direct link between them.

The relative order of the MRI-derived brain volume events were consistent across cohorts, albeit with some variance (average KTC of 0.64 ± 0.24 for MRI variables only). While the volume changes in JADNI, ARWIBO and WMHAD started with ventricular expansion and were then followed by atrophy of the temporal lobe (here, hippocampus, entorhinal, middle temporal, and fusiform gyrus), in other cohorts (ADNI, ANM, OASIS, NACC, EDSD) atrophy of the temporal lobe regions were the first detected variables of the MRI modality. The position that was taken by each respective brain region varied again among the cohorts. However, in many cases the probabilistic nature of the EBMs indicated that the order of MRI events could be interchangeable among themselves (average BC of 0.38 ± 0.21 for MRI variables only) and events occurred most probably in close temporal proximity or even simultaneously (**Figure S2**), as far as the model could discern from the data.

The position of FDG-PET, another well-established imaging biomarker measuring brain hypometabolism, was consistent in both cohorts it was measured in (ADNI, JADNI). It preceded the MRI marker changes and occurred concurrently with clinical symptoms, being placed after logical memory tests such as the LIMM and LDEL, and before assessments of executive function such as the TRABS and DIGIT.

### 3.2. Meta-sequence

To aggregate and investigate the complementary information from the base sequences in each cohort, we combined them into a single meta-sequence. Here, the position of a variable was determined based on its relative positions in all cohort sequences. All considered base-sequences and the resulting meta-sequence are presented in **Figure 2**.

**Figure 2.**
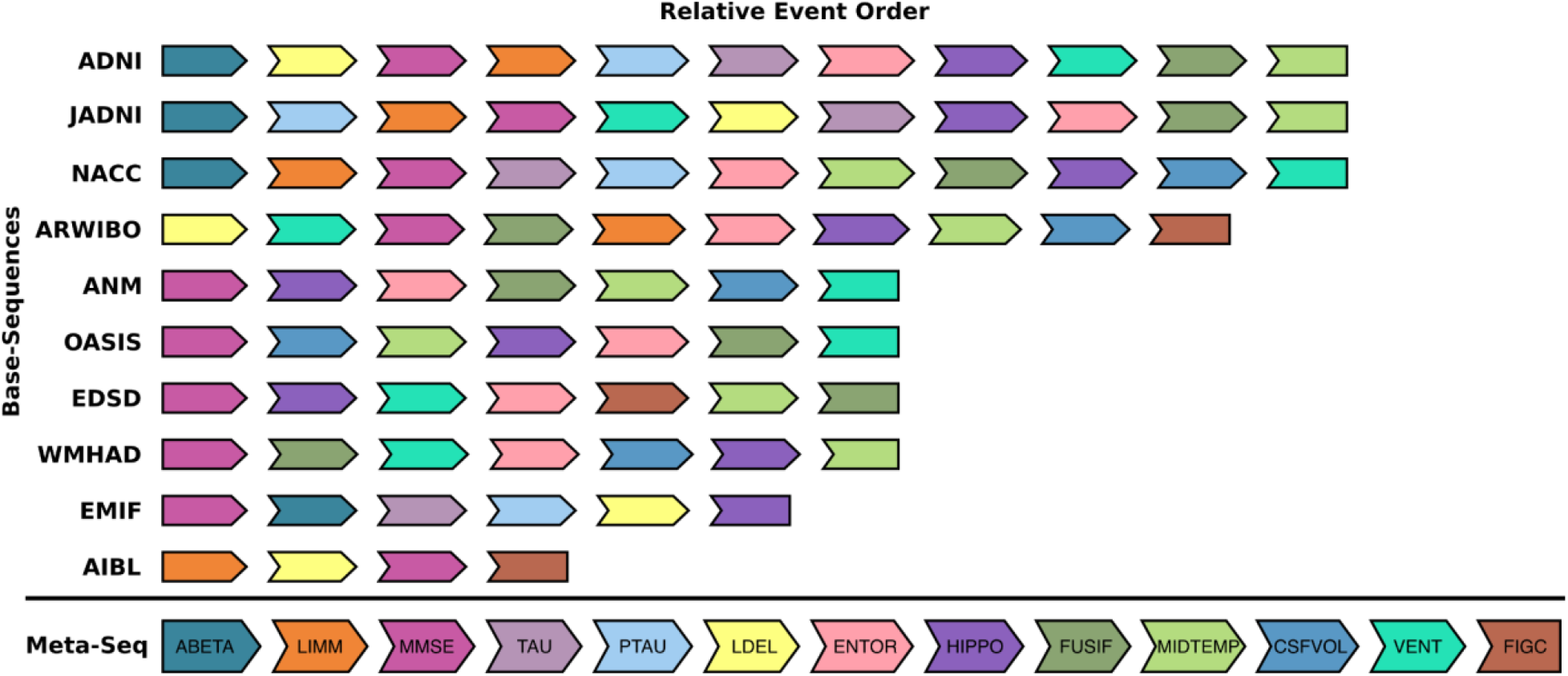
All considered base-sequences and the resulting meta-sequence. Due to only partially overlapping lists, the determining factor for an events position in the meta-sequence was not its absolute position in each base-sequence (i.e., rank 1, 2, …, 11), but its relative position to other biomarkers in the same sequence (e.g., ABETA commonly places before MMSE when they were assessed together, thus it appears before MMSE in the meta-sequence).

In the final meta-sequence, ABETA was ranked first, followed by LIMM, a clinical assessment measuring memory impairment. Although MMSE took the first rank in the majority of the base-sequences, from an algorithmic perspective, it was correctly positioned after ABETA and LIMM as they most often occurred before MMSE in cohorts where all three variables were available. In position four and five, TAU and PTAU followed respectively, again closely linked and seemingly interchangeable given their ambiguous positioning across the base-sequences. After these CSF biomarkers, the meta-model suggests that executive function (here expressed through LDEL) becomes abnormal. The later event ranks were covered by MRI markers of brain volume, starting with the temporal lobe (e.g., entorhinal cortex and hippocampus), and ending with the ventricles. The previously described ambiguity in the order of MRI regions is not reflected in the meta-sequence because the algorithm considers only the ranks, and not the uncertainty estimated by the EBM, however, it is sensible to consider MRI events as fairly interchangeable in the meta-model.

Finally, FIGC, an assessment of visual function, was positioned last which reflects its relatively stable position observed across the cohorts in which it was assessed (ARWIBO, AIBL, EDSD).

## 4. Discussion

In this work, we used EBMs to investigate AD progression across ten independent cohort studies by evaluating the concurrence of their individually derived event sequences. Furthermore, we proposed an algorithm to combine event sequences estimated from partially overlapping, and thus complementary, sets of variables into a single meta-sequence describing AD progression more comprehensively. Finally, we applied said algorithm on the ten event sequences to estimate a meta-sequence comprising 13 AD variables spanning CSF biomarkers, MRI measures and clinical assessments of cognitive and functional performance.

### 4.1. Consistent trends across cohorts’ event sequences

The derived event sequences proved to be broadly consistent across cohorts, with the most notable variability in the ordering of MRI brain volume events. This could be caused by 1) distinct statistical biases of the cohorts for example introduced through specific recruitment criteria **[21]**, 2) distinct prevalence of AD disease progression subtypes that follow different disease mechanisms **[38-40]**, or 3) mixed neuropathologies.

Inclusion and exclusion criteria of a study shape the demographic compositions of its cohort and thus can directly affect the data-driven disease progression patterns (Table S2). For instance, JADNI held a higher proportion of females compared to ADNI. Given that it has been repeatedly reported that early TAU depositioning is more prominent in female AD patients **[41-43]**, this difference might explain the earlier positioning of TAU biomarkers in JADNI’s sequence opposed to their relatively lower ranks in ADNI’s.

Previously, for example, two empirically determined AD progression subtypes called ‘hippocampal-sparing’ and ‘limbic-predominant’ were described and associated with distinct patterns of brain atrophy **[38,44]**. While structural changes in the brain start with atrophy in the medial temporal lobe (e.g., entorhinal and hippocampus) for the limbic-predominant subtype, the brain deterioration in the hippocampal-sparing subtype begins with atrophy of the frontal cortex and with the enlargement of ventricles **[45]**. Given their respective event sequences, this could indicate that OASIS, ADNI and NACC might have included more patients expressing the limbic-predominant subtype, while the hippocampal-sparing subtype was more dominant among patients from ARWIBO and JADNI.

The observed trend that global cognitive assessments like the CDRSB and MMSE placed among the first ranks in many of the event sequences, however, constituted an artifact of the diagnostic procedure. These respective assessments were used for differentiating the participants into CU and AD and hence also indirectly informed our data-driven quantification of what constitutes normal and abnormal. Therefore, observing global cognitive assessments in the early positions of a sequence should be understood more as a positive control rather than being subject to biological interpretation. In fact, previous biomarker studies have shown that both ABETA accumulation and brain atrophy can be detected before global cognitive decline **[46-49]**.

Autopsies of AD patients have shown that AD pathology hardly appears in isolation and that patients often suffer from a mixture of brain pathologies **[50]**. While most studies aim to exclude patients affected by other cognitive diseases, an AD clinical diagnosis is still mainly symptom driven and misclassification errors are possible.

### 4.2. Meta-sequence resembles AD pathology

One possibility to validate the derived meta-sequence was to evaluate its concordance with previous findings describing the temporal relationship between smaller subgroups of variables.

Previous EBM studies reported that ABETA abnormality was immediately followed by TAU and PTAU **[15,20]**, while we observed that cognitive tests placed between them in most base-sequences and, more conclusively, in the meta-sequence. However, two of these tests (LDEL and LIMM) were not included in the previous studies. In a recent study, Luo *et al*. **[51]** showed that TAU and PTAU became abnormal after ABETA and that their abnormality occurred in close temporal relationship with cognitive decline. Furthermore, there is a well-established association between cognitive decline and ABETA abnormality and abundant evidence that changes in cognition typically occur after the development of ABETA pathology **[51-53]**.

Our observation that memory function showed abnormality before brain volumes agrees with previous studies which suggested that individual-level brain atrophy rates (not assessed in our study) precede cognitive events, however, MRI-derived brain volumes become abnormal afterwards **[15]**.

In the meta-sequence, changes in MRI biomarkers were ranked after TAU and PTAU. In agreement with this, Luo *et al*. **[51]** reported that changes in MRI markers appear in close succession with TAU and PTAU. Moreover, multiple studies detected a significant correlation between TAU indices and temporal lobe atrophy **[54-56]**. Based on these studies, increases of TAU and PTAU are attributable to the deposition of neurofibrillary tangles in the temporal lobe, including the hippocampus and middle temporal. This elevated TAU predicts future brain atrophy in these regions (i.e., TAU becomes abnormal before MRI regions).

In concordance with the relative positioning of MRI biomarkers in the meta-sequence, various studies have shown that volumetric changes start with the temporal lobe areas, including the entorhinal cortex which preceded the abnormality of hippocampus and middle temporal, and further proceeds to other brain regions such as the ventricles **[57-60]**.

Finally, in agreement with a previous study **[61]** in which visual memory dysfunction was identified as one of the last stages in AD progression, the FIGC test was ranked last. The fact that it was positioned after the enlargement of ventricles further is in agreement with experimental evidence that changes in the ventricles may precede a deficit in visual memory function **[62]**. Furthermore, another EBM study **[35]** also suggested that visual processing becomes impaired after episodic memory in typical AD.

### 4.3. Limitations

The EBM estimates a single sequence, and its uncertainty, in a dataset. The uncertainty of events, and the differences across sequences estimated in distinct cohorts provides hints about possible heterogeneity of disease progression. Our proposed rank aggregation algorithm does not take the positional uncertainty into account and, like the EBM, displays the general progression pattern. To build a robust meta-sequence, each variable had to be present in at least some of the base-sequences to allow for meaningful distance calculations. Furthermore, the high amounts of missing data occurring when multiple data modalities are combined led to a substantial decrease of the number of available participants per study. This could have led to more noise in the EBM’s reference distributions.

## 5. Conclusion

In the light of the reproducibility crisis, it becomes especially vital that we look beyond single data resources, validate achieved results across multiple cohort studies, and constantly develop and evaluate data-driven methods. To this end, we revealed good consistency across data-driven event sequences derived from ten independent cohorts using EBMs. Here, only relatively minor differences in the ranking of the core features that were available in all ten cohorts were observed. In addition, our novel algorithm estimated a meta-sequence that exploits the additional information available in other variables unique to each study and thus could assemble an event sequence that is highly multimodal and more comprehensive than sequences built from single datasets. This is important for ensuring transferability of models and results across AD (sub)populations, and for improving our understanding of disease progression.

## Data Availability

De-identified data used in preparation of this article were obtained from the Alzheimer’s Disease Neuroimaging Initiative (ADNI) (https://adni.loni.usc.edu), the Australian Imaging, Biomarker and Lifestyle Flagship Study of Ageing (AIBL) database (https://aibl.csiro.au/), the European Collaboration for the Discovery of Novel Biomarkers for Alzheimer’s Disease (AddNeuroMed) (https://www.synapse.org/#!Synapse:syn4988768), Alzheimer’s Disease Repository Without Borders (ARWIBO) (https://www.neugrid2.eu/), Open Access Series of Imaging Studies (OASIS) (https://www.neugrid2.eu/), White Matter Hyperintensities in Alzheimer’s Disease (WMH-AD) (https://www.neugrid2.eu/), European Diffusion Tensor Imaging Study in Dementia (EDSD) (https://www.neugrid2.eu/), National Alzheimer’s Coordinating Center (NACC) (https://naccdata.org/), Japanese Alzheimer’s Disease Neuroimaging Initiative (JADNI) (https://humandbs.biosciencedbc.jp/en/hum0043-v1), European Medical Information Framework for Alzheimer’s Disease Multimodal Biomarker Discovery (EMIF-AD MBD) (https://emif-catalogue.eu; http://www.emif.eu/about/emif-ad).

## List of abbreviations

ADNI: The Alzheimer’s Disease Neuroimaging Initiative
JADNI: Japanese Alzheimer’s Disease Neuroimaging Initiative
AIBL: The Australian Imaging, Biomarker Lifestyle Flagship Study of Ageing
NACC: The National Alzheimer’s Coordinating Center
ANM: AddNeuroMed
EMIF-1000: European Medical Information Framework
EDSD: European DTI Study on Dementia
ARWIBO: Alzheimer’s Disease Repository Without Borders
OASIS: Open Access Series of Imaging Studies
WMHAD: White Matter Hyperintensities in Alzheimer’s Disease
CDRSB: Clinical Dementia Rating Sum of Boxes
NPI: Neuropsychiatric Inventory
LDEL: Logical Memory - Delayed Recall Total Number of Story Units Recalled
ADAS13: Alzheimer’s Disease Assessment Scale (13-items)
ADAS11: Alzheimer’s Disease Assessment Scale (11-items)
MMSE: Mini-Mental State Examination
LIMM: Logical Memory - Immediate Recall Total Number of Story Units Recalled
TRABS: Trail Making Test-B
DIGITS: Digit-Symbol Coding Test
LIDE: California Verbal Learning Test Delayed Raw Score
CATFLU: Category Fluency (animals - fruits/vegetables)
FIGC: Figure Copy
LIRE: California Verbal Learning Test Recall Raw Score
FIGR: Figure recall
STROOP: C/D Stroop Test Raw
STM: Short Term Memory
LANGU: Language
ORIENT: Perceptual Orientation
MENMA: Mental Manipulation
ATTEN: Attention
CLKS: Clock Drawing Test Total Score
EXECUTIVE: Executive Memory
LICOR: Word List Learning Trial
BNTS: Boston Naming Test Score
WAIS: Digit Symbol Substitution Test
ABETA: Amyloid-β
TAU: Total Tau
PTAU: Phosphorylated Tau (p-Tau)
ENTOR: Entorhinal Volume
HIPPO: Hippocampal Volume
FUSIF: Fusiform Volume
VENT: Ventricles Volume
MIDTEPM: MiddleTemporal Volume
CSFVOL: Accumulated CSF in the Brain
FDG: Fluorodeoxyglucose Positron Emission Tomography (FDG PET)
MRI: Magnetic resonance imaging
MCI: Mild cognitive impairment
AD: Alzheimer’s disease
CU: Cognitive unimpairment
KTC: Kendall’s tau rank correlations
EBM: Event-based model
CSF: Cerebrospinal fluid

## Declarations

## Ethics approval and consent to participate

Participants of every cohort dataset that was used in this work gave informed written consent for data collection and sharing. For more details, we refer to the provided references of each cohort, respectively.

## Consent for publication

The authors submitted the manuscript to all data owners who require manuscript approval prior to publication and acquired consent.

## Acknowledgments

We want to commend all data owners on their adherence to open science principles by sharing their data. We believe that their commitment is invaluable for AD research.

Data collection and sharing for this project were funded by the Alzheimer’s Disease Neuroimaging Initiative (ADNI; National Institutes of Health Grant U01 AG024904) and DOD ADNI (Department of Defense award number W81XWH-12-2-0012). ADNI is funded by the National Institute on Aging, the National Institute of Biomedical Imaging and Bioengineering, and through generous contributions from the following: AbbVie; Alzheimer’s Association; Alzheimer’s Drug Discovery Foundation; Araclon Biotech; BioClinica, Inc.; Biogen; Bristol-Myers Squibb Company; CereSpir, Inc.; Cogstate; Eisai Inc.; Elan Pharmaceuticals, Inc.; Eli Lilly and Company; EuroImmun; F.Hoffmann-La Roche Ltd and its affiliated company Genentech, Inc.; Fujirebio; GE Healthcare; IXICO Ltd; Janssen Alzheimer Immunotherapy Research Development, LLC; Johnson Johnson Pharmaceutical Research Development LLC; Lumosity; Lundbeck; Merck Co., Inc.; Meso Scale Diagnostics, LLC; NeuroRx Research; Neurotrack Technologies; Novartis Pharmaceuticals Corporation; Pfizer Inc.; Piramal Imaging; Servier; Takeda Pharmaceutical Company; and Transition Therapeutics. The Canadian Institutes of Health Research is providing funds to support ADNI clinical sites in Canada. Private-sector contributions are facilitated by the Foundation for the National Institutes of Health (www.fnih.org). The grantee organization is the Northern California Institute for Research and Education, and the study is coordinated by the Alzheimer’s Therapeutic Research Institute at the University of Southern California. ADNI data are disseminated by the Laboratory for NeuroImaging at the University of Southern California.

Data collection and sharing of ARWIBO was supported by the Italian Ministry of Health, under the following grant agreements: Ricerca Corrente IRCCS Fatebenefratelli, Linea di Ricerca 2; Progetto Finalizzato Strategico 2000-2001 “Archivio normativo italiano di morfometria cerebrale con risonanza magnetica (età 40+)”; Progetto Finalizzato Strategico 2000-2001 “Decadimento cognitivo lieve non dementigeno: stadio preclinico di malattia di Alzheimer e demenza vascolare. Caratterizzazione clinica, strumentale, genetica e neurobiologica e sviluppo di criteri diagnostici utilizzabili nella realtà nazionale,”; Progetto Finalizzata 2002 “Sviluppo di indicatori di danno cerebrovascolare clinicamente significativo alla risonanza magnetica strutturale”; Progetto Fondazione CARIPLO 2005-2007 “Geni di suscettibilità per gli endofenotipi associati a malattie psichiatriche e dementigene”; “Fitness and Solidarietà”; and anonymous donors.

DNI was supported by the following grants: Translational Research Promotion Project from the New Energy and Industrial Technology Development Organization of Japan; Research on Dementia, Health Labor Sciences Research Grant; Life Science Database Integration Project of Japan Science and Technology Agency; Research Association of Biotechnology (contributed by Astellas Pharma Inc., Bristol-Myers Squibb, Daiichi-Sankyo, Eisai, Eli Lilly and Company, Merck-Banyu, Mitsubishi Tanabe Pharma, Pfizer Inc., Shionogi Co., Ltd., Sumitomo Dainippon, and Takeda Pharmaceutical Company), Japan, and a grant from an anonymous foundation.

The NACC database is funded by NIA/NIH Grant U01 AG016976. NACC data are contributed by the NIA-funded ADCs: P30 AG019610 (PI Eric Reiman, MD), P30 AG013846 (PI Neil Kowall, MD), P30 AG062428-01 (PI James Leverenz, MD) P50 AG008702 (PI Scott Small, MD), P50 AG025688 (PI Allan Levey, MD, PhD), P50 AG047266 (PI Todd Golde, MD, PhD), P30 AG010133 (PI Andrew Saykin, PsyD), P50 AG005146 (PI Marilyn Albert, PhD), P30 AG062421-01 (PI Bradley Hyman, MD, PhD), P30 AG062422-01 (PI Ronald Petersen, MD, PhD), P50 AG005138 (PI Mary Sano, PhD), P30 AG008051 (PI Thomas Wisniewski, MD), P30 AG013854 (PI Robert Vassar, PhD), P30 AG008017 (PI Jeffrey Kaye, MD), P30 AG010161 (PI David Bennett, MD), P50 AG047366 (PI Victor Henderson, MD, MS), P30 AG010129 (PI Charles DeCarli, MD), P50 AG016573 (PI Frank LaFerla, PhD), P30 AG062429-01(PI James Brewer, MD, PhD), P50 AG023501 (PI Bruce Miller, MD), P30 AG035982 (PI Russell Swerdlow, MD), P30 AG028383 (PI Linda Van Eldik, PhD), P30 AG053760 (PI Henry Paulson, MD, PhD), P30 AG010124 (PI John Trojanowski, MD, PhD), P50 AG005133 (PI Oscar Lopez, MD), P50 AG005142 (PI Helena Chui, MD), P30 AG012300 (PI Roger Rosenberg, MD), P30 AG049638 (PI Suzanne Craft, PhD), P50 AG005136 (PI Thomas Grabowski, MD), P30 AG062715-01 (PI Sanjay Asthana, MD, FRCP), P50 AG005681 (PI John Morris, MD), P50 AG047270 (PI Stephen Strittmatter, MD, PhD).

## Availability of data and materials

De-identified data used in preparation of this article were obtained from the Alzheimer’s Disease Neuroimaging Initiative (ADNI) (https://adni.loni.usc.edu), the Australian Imaging, Biomarker and Lifestyle Flagship Study of Ageing (AIBL) database (https://aibl.csiro.au/), the European Collaboration for the Discovery of Novel Biomarkers for Alzheimer’s Disease (AddNeuroMed) (https://www.synapse.org/#!Synapse:syn4988768), Alzheimer’s Disease Repository Without Borders (ARWIBO) (https://www.neugrid2.eu/), Open Access Series of Imaging Studies (OASIS) (https://www.neugrid2.eu/), White Matter Hyperintensities in Alzheimer’s Disease (WMH-AD) (https://www.neugrid2.eu/), European Diffusion Tensor Imaging Study in Dementia (EDSD) (https://www.neugrid2.eu/), National Alzheimer’s Coordinating Center (NACC) (https://naccdata.org/), Japanese Alzheimer’s Disease Neuroimaging Initiative (JADNI) (https://humandbs.biosciencedbc.jp/en/hum0043-v1), European Medical Information Framework for Alzheimer’s Disease Multimodal Biomarker Discovery (EMIF-AD MBD) (https://emif-catalogue.eu; http://www.emif.eu/about/emif-ad). The authors had no special access privileges others would not have to the data obtained from these resources.

## Author contributions

CB conceived and designed the study. YS and CB collected the datasets. NPO and SGK implemented the methods. SGK prepared the data and ran the experiments. CB and SGK drafted the manuscript. YS, NPO, and MHA revised the manuscript. CB and NPO supervised the work. MHA acquired the primary funding.

## References

1. Jack, C. R., Jr, Bennett, D. A., Blennow, K., Carrillo, M. C., Dunn, B., Haeberlein, S. B., et al. NIA-AA Research Framework: Toward a biological definition of Alzheimer’s disease. Alzheimers Dement 2018; 14(4), 535–562. https://doi.org/10.1016/j.jalz.2018.02.018

2. DeTure, M. A., Dickson, D. W. The neuropathological diagnosis of Alzheimer’s disease. Mol neurodegeneration 2019; 14(1), 32. https://doi.org/10.1186/s13024-019-0333-5

3. Blennow, K., Zetterberg, H. Biomarkers for Alzheimer’s disease: current status and prospects for the future. Intern Med 2018; 284(6), 643–663. https://doi.org/10.1111/joim.12816

4. Lovestone, S., Francis, P., Kloszewska, I., Mecocci, P., Simmons, A., Soininen, H., et al. AddNeuroMed--the European collaboration for the discovery of novel biomarkers for Alzheimer’s disease. Ann N Y Acad Sci 2009; 1180, 36–46. https://doi.org/10.1111/j.1749-6632.2009.05064.x

5. Lorenzi, M., Filippone, M., Frisoni, G. B., Alexander, D. C., Ourselin, S., Alzheimer’s Disease Neuroimaging Initiative. Probabilistic disease progression modeling to characterize diagnostic uncertainty: Application to staging and prediction in Alzheimer’s disease. 2019; NeuroImage, 190, 56–68. https://doi.org/10.1016/j.neuroimage.2017.08.059

6. Jedynak, B. M., Lang, A., Liu, B., Katz, E., Zhang, Y., Wyman, B. T., et al. A computational neurodegenerative disease progression score: method and results with the Alzheimer’s disease Neuroimaging Initiative cohort. 2012; NeuroImage, 63(3), 1478–1486. https://doi.org/10.1016/j.neuroimage.2012.07.059

7. Yang, E., Farnum, M., Lobanov, V., Schultz, T., Verbeeck, R., Raghavan, N., Samtani, M. N., Novak, G., Narayan, V., DiBernardo, A., Alzheimer’s Disease Neuroimaging Initiative. Quantifying the pathophysiological timeline of Alzheimer’s disease. J Alzheimers Dis 2011; 26(4), 745–753. https://doi.org/10.3233/JAD-2011-110551

8. Delor, I., Charoin, J. E., Gieschke, R., Retout, S., Jacqmin, P. Modeling Alzheimer’s Disease Progression Using Disease Onset Time and Disease Trajectory Concepts Applied to CDR-SOB Scores From ADNI. CPT pharmacometrics systems pharmacology 2013; 2(10), e78. https://doi.org/10.1038/psp.2013.54

9. Villemagne, V. L., Burnham, S., Bourgeat, P., Brown, B., Ellis, K. A., Salvado, O., et al. Amyloid β deposition, neurodegeneration, and cognitive decline in sporadic Alzheimer’s disease: a prospective cohort study. Lancet Neurol 2013; 12(4), 357–367. https://doi.org/10.1016/S1474-4422(13)70044-9

10. Donohue, M. C., Jacqmin-Gadda, H., Le Goff, M., Thomas, R. G., Raman, R., Gamst, A. et al. Estimating long-term multivariate progression from short-term data. Alzheimers dement 2014, 10(5 Suppl), S400–S410. https://doi.org/10.1016/j.jalz.2013.10.003

11. Dekker, I., Schoonheim, M. M., Venkatraghavan, V., Eijlers, A., Brouwer, I., Bron, E. E., et al. The sequence of structural, functional and cognitive changes in multiple sclerosis. NeuroImage 2021; Clinical, 29, 102550. https://doi.org/10.1016/j.nicl.2020.102550

12. Oxtoby, N. P., Leyland, L. A., Aksman, L. M., Thomas, G., Bunting, E. L., Wijeratne, P. et al. Sequence of clinical and neurodegeneration events in Parkinson’s disease progression. Brain 2021; 144(3), 975–988. https://doi.org/10.1093/brain/awaa461

13. Fonteijn, H. M., Modat, M., Clarkson, M. J., Barnes, J., Lehmann, M., Hobbs, N. Z., et al. An event-based model for disease progression and its application in familial Alzheimer’s disease and Huntington’s disease. NeuroImage 2012; 60(3), 1880–1889. https://doi.org/10.1016/j.neuroimage.2012.01.062

14. Wijeratne, P. A., Young, A. L., Oxtoby, N. P., Marinescu, R. V., Firth, N. C., Johnson, E. et al. An image-based model of brain volume biomarker changes in Huntington’s disease. Ann Clin Transl Neurol 2018; 5(5), 570–582. https://doi.org/10.1002/acn3.558

15. Young, A. L., Oxtoby, N. P., Daga, P., Cash, D. M., Fox, N. C., Ourselin, S., Schott, J. et al. A data-driven model of biomarker changes in sporadic Alzheimer’s disease. Brain 2014; 137(Pt 9), 2564–2577. https://doi.org/10.1093/brain/awu176

16. Young, A. L., Marinescu, R. V., Oxtoby, N. P., Bocchetta, M., Yong, K., Firth, N. C., et al. Uncovering the heterogeneity and temporal complexity of neurodegenerative diseases with Subtype and Stage Inference. Nat Commun 2018; 9(1), 4273. https://doi.org/10.1038/s41467-018-05892-0

17. Mueller, S. G., Weiner, M. W., Thal, L. J., Petersen, R. C., Jack, C. R., Jagust, W., et al. Ways toward an early diagnosis in Alzheimer’s disease: the Alzheimer’s Disease Neuroimaging Initiative (ADNI). Alzheimers Dement 2005; 1(1), 55–66.

18. Solomon, A., Kivipelto, M., Molinuevo, J. L., Tom, B., Ritchie, C. W.. European prevention of Alzheimer’s dementia longitudinal cohort study (EPAD LCS): study protocol. Prev Alzheimers Dis 2018, 8(12), e021017.

19. Oxtoby, N. P., Young, A. L., Cash, D. M., Benzinger, T., Fagan, A. M., Morris, J. C., et al. Data-driven models of dominantly-inherited Alzheimer’s disease progression. Brain 2018; 141(5), 1529–1544. https://doi.org/10.1093/brain/awy050

20. Archetti, D., Ingala, S., Venkatraghavan, V., Wottschel, V., Young, A. L., Bellio, M., et al. Multi-study validation of data-driven disease progression models to characterize evolution of biomarkers in Alzheimer’s disease. NeuroImage 2019; 24, 101954. https://doi.org/10.1016/j.nicl.2019.101954

21. Birkenbihl, C., Salimi, Y., Fröhlich, H., Japanese Alzheimer’s Disease Neuroimaging Initiative, Alzheimer’s Disease Neuroimaging Initiative. Unraveling the heterogeneity in Alzheimer’s disease progression across multiple cohorts and the implications for data-driven disease modeling. Alzheimers dement 2021; https://doi.org/10.1002/alz.12387

22. Birkenbihl, C., Emon, M. A., Vrooman, H., Westwood, S., Lovestone, S., et al. Differences in cohort study data affect external validation of artificial intelligence models for predictive diagnostics of dementia - lessons for translation into clinical practice. The EPMA 2020; 11(3), 367–376. https://doi.org/10.1007/s13167-020-00216-z

23. Salimi, Y., Domingo-Fernandez, D., Bobis-Alvarez, C., Hofmann-Apitius, M., Vasculature, I., Birkenbihl, C. et al. ADataViewer: Exploring Semantically Harmonized Alzheimer’s Disease Cohort Datasets. medRxiv 2021.

24. McKhann, G., Drachman, D., Folstein, M., Katzman, R., Price, D., Stadlan, E. M. Clinical diagnosis of Alzheimer’s disease: report of the NINCDS-ADRDA Work Group under the auspices of Department of Health and Human Services Task Force on Alzheimer’s Disease. Neurology 1984; 34(7), 939–944. https://doi.org/10.1212/wnl.34.7.939

25. Iwatsubo, T. Japanese Alzheimer’s Disease Neuroimaging Initiative: present status and future. Alzheimer Dement 2010; 6(3), 297–299. https://doi.org/10.1016/j.jalz.2010.03.011

26. Ellis, K. A., Bush, A. I., Darby, D., De Fazio, D., Foster, J., Hudson, P, et al. The Australian Imaging, Biomarkers and Lifestyle (AIBL) study of aging: methodology and baseline characteristics of 1112 individuals recruited for a longitudinal study of Alzheimer’s disease. Int Psychogeriatr 2009; 21(4), 672–687. https://doi.org/10.1017/S1041610209009405

27. Besser, L., Kukull, W., Knopman, D. S., Chui, H., Galasko, D., Weintraub, S. et al. Version 3 of the National Alzheimer’s Coordinating Center’s Uniform Data Set. Alzheimer Dis Assoc Disord 2018; 32(4):351–358. https://doi.org/10.1097/WAD.0000000000000279

28. Birkenbihl, C., Westwood, S., Shi, L., Nevado-Holgado, A., Westman, E., Lovestone, S., et al. ANMerge: a comprehensive and accessible Alzheimer’s disease patient-level dataset. J Alzheimers Dis 2021; 79(1):423–431. https://doi.org/10.3233/JAD-200948.

29. Bos, I., Vos, S., Vandenberghe, R., Scheltens, P., Engelborghs, S., Frisoni, G.,et al The EMIF-AD Multimodal Biomarker Discovery study: design, methods and cohort characteristics. Alzheimers Res Ther (2018); 10(1), 1–9. https://doi.org/10.1186/s13195-018-0396-5

30. Brueggen, K., Grothe, M. J., Dyrba, M., Fellgiebel, A., Fischer, F., Filippi, M. et al. The European DTI Study on Dementia—a multicenter DTI and MRI study on Alzheimer’s disease and mild cognitive impairment. NeuroImage 2017; 144, 305–308. https://doi.org/10.1016/j.neuroimage.2016.03.067

31. Frisoni, G. B., Prestia, A., Zanetti, O., Galluzzi, S., Romano, M., Cotelli, M. et al. Markers of Alzheimer’s disease in a population attending a memory clinic. Alzheimers Dement 2009; 5(4), 307–317. https://doi.org/10.1016/j.jalz.2009.04.1235

32. Marcus, D. S., Wang, T. H., Parker, J., Csernansky, J. G., Morris, J. C., Buckner, R. L. (2007). Open Access Series of Imaging Studies (OASIS): cross-sectional MRI data in young, middle aged, nondemented, and demented older adults. J Cogn Neurosci 2007; 19(9), 1498–1507. https://doi.org/10.1162/jocn.2007.19.9.1498

33. Marcus, D. S., Fotenos, A. F., Csernansky, J. G., Morris, J. C., Buckner, R. L. Open access series of imaging studies: longitudinal MRI data in nondemented and demented older adults. J Cogn Neurosci 2010; 22(12), 2677–2684. https://doi.org/10.1162/jocn.2009.21407

34. Damulina, A., Pirpamer, L., Seiler, S., Benke, T., Dal-Bianco, P., Ransmayr, G., et al. White matter hyperintensities in Alzheimer’s disease: a lesion probability mapping study. J Alzheimers Dis 2019; 68(2), 789–796. https://doi.org/10.3233/JAD-180982

35. Firth, N. C., Primativo, S., Brotherhood, E., Young, A. L., Yong, K., Crutch, S. J., et al. Sequences of cognitive decline in typical Alzheimer’s disease and posterior cortical atrophy estimated using a novel event-based model of disease progression. Alzheimers demen 2020; 16(7), 965–973. https://doi.org/10.1002/alz.12083

36. DeConde, R. P., Hawley, S., Falcon, S., Clegg, N., Knudsen, B., Etzioni, R. Combining results of microarray experiments: a rank aggregation approach. Stat Appl Genet Mol Biol 2006; 5, Article15. https://doi.org/10.2202/1544-6115.1204

37. Lin, S., Ding, J. Integration of ranked lists via cross entropy Monte Carlo with applications to mRNA and microRNA Studies. Biometrics 2009; 65(1), 9–18. https://doi.org/10.1111/j.1541-0420.2008.01044.x

38. Ferreira, D., Nordberg, A., Westman, E. Biological subtypes of Alzheimer disease: A systematic review and meta-analysis. Neurology 2020; 94(10), 436–448. https://doi.org/10.1212/WNL.0000000000009058

39. Whitwell, J. L., Jack Jr, C. R., Przybelski, S. A., Parisi, J. E., Senjem, M. L., Boeve, B. F., et al. Temporoparietal atrophy: a marker of AD pathology independent of clinical diagnosis. Neurobiol Aging 2011; 32(9), 1531–1541, https://doi.org/10.1016/j.neurobiolaging.2009.10.012

40. Piaceri, I., Nacmias, B., Sorbi, S. (2013). Genetics of familial and sporadic Alzheimer’s disease. Front Biosci 2013; 5 (1), 167–177. https://doi.org/10.2741/e605

41. Smith, R., Strandberg, O., Mattsson-Carlgren, N., Leuzy, A., Palmqvist, S., Pontecorvo, et al. The accumulation rate of tau aggregates is higher in females and younger amyloid-positive subjects. Brain 2020; 143(12), 3805–3815. https://doi.org/10.1093/brain/awaa327

42. Deming, Y., Dumitrescu, L., Barnes, L. L., Thambisetty, M., Kunkle, B., Gifford, K. A., et al. Sex-specific genetic predictors of Alzheimer’s disease biomarkers. Acta neuropathol 2018, 136(6), 857–872. https://doi.org/10.1007/s00401-018-1881-4

43. Sundermann, E. E., Panizzon, M. S., Chen, X., Andrews, M., Galasko, D., Banks, S. J., et al. Sex differences in Alzheimer’s-related Tau biomarkers and a mediating effect of testosterone. Biol Sex Differ 2020; 11(1), 33. https://doi.org/10.1186/s13293-020-00310-x

44. Ferreira, D., Pereira, J. B., Volpe, G., Westman, E. Subtypes of Alzheimer’s Disease Display Distinct Network Abnormalities Extending Beyond Their Pattern of Brain Atrophy. Front Neurol. 2019; 10, 524. https://doi.org/10.3389/fneur.2019.00524

45. Ferreira, D., Verhagen, C., Hernández-Cabrera, J. A., Cavallin, L., Guo, C. J., Ekman, U., et al. Distinct subtypes of Alzheimer’s disease based on patterns of brain atrophy: longitudinal trajectories and clinical applications. Sci Rep 2017; 7, 46263. https://doi.org/10.1038/srep46263

46. Iturria-Medina, Y., Sotero, R. C., Toussaint, P. J., Mateos-Pérez, J. M., Evans, A. C., et al. Early role of vascular dysregulation on late-onset Alzheimer’s disease based on multifactorial data-driven analysis. Nat. Commun 2016; 7, 11934. https://doi.org/10.1038/ncomms11934

47. Chen, G., Shu, H., Chen, G., Ward, B. D., Antuono, P. G., Zhang, Z., Li, S. J. et al. Staging Alzheimer’s Disease Risk by Sequencing Brain Function and Structure, Cerebrospinal Fluid, and Cognition Biomarkers. J Alzheimers Dis 2016; 54(3), 983–993. https://doi.org/10.3233/JAD-160537

48. Mormino, E. C., Kluth, J. T., Madison, C. M., Rabinovici, G. D., Baker, S. L., Miller, B. et al. Episodic memory loss is related to hippocampal-mediated beta-amyloid deposition in elderly subjects. Brain 2009; 132(Pt 5), 1310–1323. https://doi.org/10.1093/brain/awn3

49. Wang, F., Gordon, B. A., Ryman, D. C., Ma, S., Xiong, C., Hassenstab, J., Goate, A., et al. Cerebral amyloidosis associated with cognitive decline in autosomal dominant Alzheimer disease. Neurology 2015; 85(9), 790–798. https://doi.org/10.1212/WNL.0000000000001903

50. Schneider, J. A., Arvanitakis, Z., Bang, W., Bennett, D. A. Mixed brain pathologies account for most dementia cases in community-dwelling older persons. Neurology 2007; 69(24), 2197–2204. https://doi.org/10.1212/01.wnl.0000271090.28148.24

51. Luo, J., Agboola, F., Grant, E., Masters, C. L., Albert, M. S., Johnson, S. C., et al. Sequence of Alzheimer disease biomarker changes in cognitively normal adults: A cross-sectional study. Neurology 2020, 95(23), e3104–e3116. https://doi.org/10.1212/WNL.0000000000010747

52. Lim, Y. Y., Ellis, K. A., Harrington, K., Pietrzak, R. H., Gale, J., Ames, D., et al. Cognitive decline in adults with amnestic mild cognitive impairment and high amyloid-β: prodromal Alzheimer’s disease? J Alzheimers Dis 2013; 33(4), 1167–1176. https://doi.org/10.3233/JAD-121771

53. Ellis, K. A., Lim, Y. Y., Harrington, K., Ames, D., Bush, A. I., Darby, D., et al. Decline in cognitive function over 18 months in healthy older adults with high amyloid-β. J Alzheimers Dis 2013; 34(4), 861–871. https://doi.org/10.3233/JAD-122170

54. Herukka, S. K., Pennanen, C., Soininen, H., Pirttilä, T. CSF Abeta42, tau and phosphorylated tau correlate with medial temporal lobe atrophy.J Alzheimers Dis 2008; 14(1), 51–57. https://doi.org/10.3233/jad-2008-14105

55. Granadillo, E., Paholpak, P., Mendez, M. F., Teng, E. Visual Ratings of Medial Temporal Lobe Atrophy Correlate with CSF Tau Indices in Clinical Variants of Early-Onset Alzheimer Disease. Dement Geriatr Cogn Disord 2017; 44(1-2), 45–54. https://doi.org/10.1159/000477718

56. Bouwman, F. H., Schoonenboom, S. N., van der Flier, W. M., van Elk, E. J., Kok, A., Barkhof, et al. CSF biomarkers and medial temporal lobe atrophy predict dementia in mild cognitive impairment. Neurobiolaging 2007; 28(7), 1070–1074. https://doi.org/10.1016/j.neurobiolaging.2006.05.006

57. Younes, L., Albert, M., Miller, M. I., BIOCARD Research Team. Inferring changepoint times of medial temporal lobe morphometric change in preclinical Alzheimer’s disease. NeuroImage 2014; Clinical, 5, 178–187. https://doi.org/10.1016/j.nicl.2014.04.009

58. Coupé, P., Manjón, J. V., Lanuza, E., Catheline, G. (2019). Lifespan Changes of the Human Brain In Alzheimer’s Disease. Sci Rep 2019; 9(1), 3998. https://doi.org/10.1038/s41598-019-39809-8

59. Scahill, R. I., Schott, J. M., Stevens, J. M., Rossor, M. N., Fox, N. C. Mapping the evolution of regional atrophy in Alzheimer’s disease: unbiased analysis of fluid-registered serial MRI. Proc Natl Acad Sci U S A 2002; 99(7), 4703–4707. https://doi.org/10.1073/pnas.052587399

60. Braak, H., Braak, E. (1991). Neuropathological stageing of Alzheimer-related changes. Acta neuropathol 1991; 82(4), 239–259. https://doi.org/10.1007/BF00308809

61. Storey, E., Slavin, M. J., Kinsella, G. J. Patterns of cognitive impairment in Alzheimer’s disease: assessment and differential diagnosis. Front Biosci 2002; 7, e155–e184. https://doi.org/10.2741/A914

62. Breteler, M. M., van Amerongen, N. M., van Swieten, J. C., Claus, J. J., Grobbee, D. E., van Gijn, et al. Cognitive correlates of ventricular enlargement and cerebral white matter lesions on magnetic resonance imaging. The Rotterdam Study. Stroke 1994; 25(6), 1109–1115. https://doi.org/10.1161/01.str.25.6.1109

