## Supplementary files for "Comparison and aggregation of event sequences across ten cohorts to describe the consensus biomarker evolution in Alzheimer’s disease"

### Supplementary File

#### Data preprocessing

While in some of the cohorts brain volumes were calculated as a sum of the two respective hemispheres, in others they were measured individually per hemisphere and thus we had to sum them up to make the measurements consistent across all cohorts. In addition, the brain region volumes of individual cohorts were normalized across the subjects based on their whole brain volume to correct the variations of head size among individuals by dividing the regional volumes through the intracranial volume.

#### EBM

While prior versions of EBMs mainly integrated parametric mixture models (i.e. Gaussian mixture models; GMM) **[1-3]**, we leveraged its latest installment which incorporates nonparametric mixture models by employing a kernel density estimation (KDE) to determine the probability density function **[4]**. KDE estimates the probability density of independent and identically distributed samples ($x_1$, $x_2$, …, $x_n$) drawn from a distribution with an unknown density by

$$\widehat{f}(x) = \frac{1}{Nh}\sum_{j=1}^{N} K\left(\frac{x-x_j}{h}\right) \qquad \textit{Equation 2}$$

where $K$ and $h$ are kernel function and bandwidth, respectively **[5]**. The kernel we used was Gaussian and the bandwidth for estimating the components of the mixture models was determined by Scott's normal reference rule **[6]**.

#### Meta-sequence generating algorithm

Our proposed method for generating a meta-sequence from multiple, complementary base sequences represents an algorithm addressing the rank aggregation of partial lists **[7]**. It essentially solves an optimization problem (i.e., finding the meta-sequence with the smallest distance to all base-sequences) by combining an exhaustive search for the initial $k$-length starting sequence with a greedy search procedure adding missing variables into the respective position in the sequence where the average distance of the altered sequence to all base-sequences remains minimal. The reason for combining these two steps lies in the combinatorial explosion of the search space (i.e., the set of all theoretically possible meta-sequences) when including an increasing number of variables. Therefore, an exhaustive search is often computationally infeasible and heuristic approaches have to be considered instead. In such cases, rank aggregation approaches often rely on Monte Carlo sampling to test a set of random meta-sequences and then opt for the one with the lowest distance **[8]**. However, we found that these approaches often end with suboptimal meta-sequences for large search spaces and that the proposed approach mixing an exhaustive search for a starting sequence with subsequent greedy refinements leads to more robust and plausible results, both biologically and in comparison to the base sequences. The python code for running this algorithm can be found under (https://github.com/sepehrgolriz/EBM-MultiCohort).

**Meta-sequence generation algorithm**

The following pseudocode outlines the algorithm used to generate a meta-sequence out of ten independent event sequences

**Require:**

Set of all independent event sequences coming from cohorts, $\{ S \mid s \in S \}$
Set of all individual events coming from all cohort event sequences, $\{ E \mid e \in E \}$
Set of pattern sequences coming from permutation of all individual events, $\{ P \mid p \in P \}$

**function** CALCUALTE DISTANCE BETWEEN $(P, S)$
  **for all** $p \in P$ **do**
    **for all** $s \in S$ **do**
      Find all common events between $s$, $p$, $\{ CE \mid ce \in CE \}$
      **if** $CE \neq 0$ **then**
        **for** $ce \in CE$ **do**
          $ce_p$ = position of $ce$ in $p$
          $ce_s$ = position of $ce$ in $s$
          d = $| ce_p - ce_s |$
          Set of all distances between each of $ce$ in $s$, $p$, $\{ D \mid d \in D \}$
      Distances between p,s $\{ D(p,s) \mid d(p,s) \in D(p,s),\ D(p,s) = \frac{1}{N} \sum_{j=0}^{N} d(p,s) \}$
    Distances between p,S $\left\{ D(p,S) \,\middle|\, d(p,S) \in D(p,S),\ D(p,S) = \frac{1}{N} \sum_{j=0}^{N} d(p,S) \right\}$.
  $\Longrightarrow$ D(p, S) Set of pattern sequences with their corresponding average distance with cohorts
**return** D(p, S)

**Init**$_{(p)}$ = **min** D(p, S)

**function** ADDING REMAIND FEATURES (**Init(p)**, $E$, $S$)
  **for all** $e \in E$ **do**
  **if** $e$ **not in** $Init(p)$ **do**
    **for all positions *(i)* in** $Init(p)$) **do**
      Distances between $Init(p)$,s $\{D(Init(p), s) \mid d(Init(p), s) \in D(Init(p), s), D(Init(p), s)\ \frac{1}{N} \sum_{j=0}^{N} d(Init(p), s)$
  Distances between $Init(p)$,S$\left\{ D(Init(p), S) \,\middle|\, d(Init(p), S) \in D(Init(p), S),\ D(p, S) = \frac{1}{N} \sum_{j=0}^{N} d(Init(p), S) \right\}$

$\Longrightarrow$ **Init**$_{(p)}$**. insert *(e,i)*** Insert the event in the position (*i*) which the distance between the *Init(p)* and all other event sequences is minimum
  **return Init**$_{(p)}$

**Figure S1.** Proposed algorithm for determining a meta-sequence from multiple potentially only partially overlapping base sequences.

#### Handling partially overlapping lists

Distance calculations have to be performed in the same mathematical space which, in this case, is defined by the variables in the sequences to be compared. Calculating the distance between two sequences which share the same variables is therefore straight forward. However, since individual base-sequences are often only partially overlapping, such distance calculations are impeded. There are two solutions for this problem: 1) penalizing the absence of variables in either sequence such that the distance increases with a higher number of uncommon variables, or 2) ignore variables that are only present in one of the sequences when calculating the distance. In the context of clinical cohort data, whether a specific variable was assessed depends on the study's goals and funding and, as such, its absence does seldomly hold biological meaning. Therefore, in this case, penalizing the absence of variables would bias the constructed meta-sequence.

#### Distance metrics

Depending on the focus of the study, different distance metrics can be used in the proposed algorithm. Intuitive choices are Spearman's footrule distance or Kendall's tau. The former takes the magnitude of the rank differences into account, while the latter is only counting how many rank discrepancies are found between two compared sequences, ignoring their specific position. A decision on which metric should be used depends on the emphasis of the study. In this study, we used Spearman's footrule distance because it takes the absolute difference in positions of variables into account which should be informative in our biomedical context.

#### Supplementary Table

| Cohort | Memory | Executive | Language | Visuospatial | Global cognitive |
|---|---|---|---|---|---|
| ADNI | LIMM<br>LDEL | DIGIT<br>TRABS | - | - | MMSE<br>CDRSB<br>ADAS11<br>ADAS13 |
| JADNI | LIMM<br>LDEL | DIGIT<br>TRABS | - | - | MMSE<br>CDRSB<br>ADAS11<br>ADAS13 |
| NACC | LIMM | WAIS | BNTS<br>CATFLU | - | MMSE |
| AILB | LIMM<br>LDEL | WAIS<br>STROOP | LIRE<br>LIDE<br>LICOR<br>CATFLU | FIGC<br>FIGR | MMSE |
| EMIF | LIMM<br>LDEL | EXECUTIVE | LANG | | MMSE |
| ANM | - | - | - | - | MMSE |
| ARWIBO | LIMM<br>LDEL | - | - | FIGC | MMSE |
| OASIS | - | - | - | - | MMSE |
| EDSD | - | - | LIRE<br>LICOR<br>BNTS | FIGC<br>FIGR<br>CLKS | MMSE |
| WMAHD | STM | - | CATFLU | CLKS | MMSE |

**Table S1.** Cohort-specific cognitive tests composing each cognitive domain score.

| Cohort | Age | Education | Female % | APOE4% |
|---|---|---|---|---|
| ADNI | 75±6 | 16±2 | 49 | 50 |
| JADNI | 70±6 | 12±3 | 55 | 62 |
| ARWIBO | 59±13 | 8±2 | 31 | 65 |
| OASIS | 52±12 | 5±1 | 39 | - |
| EMIF | 67±7 | 12±3 | 50 | 51 |
| ANM | 59±6 | 11±3 | 46 | 58 |
| AIBL | 58±6 | 12±2 | 48 | 56 |
| EDSD | 72±6 | 12±3 | 50 | 52 |
| WMHAD | 74±8 | 8±2 | 48 | 56 |
| NACC | 65±10 | 16±2 | 55 | 55 |

**Table S2.** The table above summarizes the demographic characteristics of each cohort.

| | AIBL | JADNI | ANM | WMHAD | ARWIBO | EMIF | OASIS | ADNI | EDSD | NACC |
|---|---|---|---|---|---|---|---|---|---|---|
| **AIBL** | 1 | 0.54 | - | - | 0.72 | 0.54 | - | 0.81 | 0.59 | 0.71 |
| **JADNI** | 0.54 | 1 | 0.69 | 0.68 | 0.64 | 0.84 | 0.71 | 0.68 | 0.86 | 0.75 |
| **ANM** | - | 0.69 | 1 | 0.78 | 0.67 | - | 0.51 | 0.64 | 0.79 | 0.61 |
| **WMHAD** | - | 0.68 | 0.78 | 1 | 0.83 | - | 0.63 | 0.91 | 0.91 | 0.68 |
| **ARWIBO** | 0.72 | 0.64 | 0.67 | 0.83 | 1 | 1 | 0.83 | 0.79 | 0.69 | 0.86 |
| **EMIF** | 0.54 | 0.84 | - | - | 1 | 1 | - | 0.79 | 0.69 | 0.86 |
| **OASIS** | - | 0.71 | 0.51 | 0.63 | 0.83 | - | 1 | 0.84 | - | 0.84 |
| **ADNI** | 0.81 | 0.68 | 0.64 | 0.91 | 0.79 | 0.84 | 0.75 | 1 | 0.90 | 0.69 |
| **EDSD** | 0.59 | 0.86 | 0.79 | 0.91 | 0.59 | - | 0.76 | 0.90 | 1 | 0.83 |
| **NACC** | 0.71 | 0.75 | 0.61 | 0.68 | 0.86 | 0.91 | 0.84 | 0.69 | 0.83 | 1 |

**Table S3.** Pairwise Kendall’s tau rank correlation coefficients

### Supplementary Figure

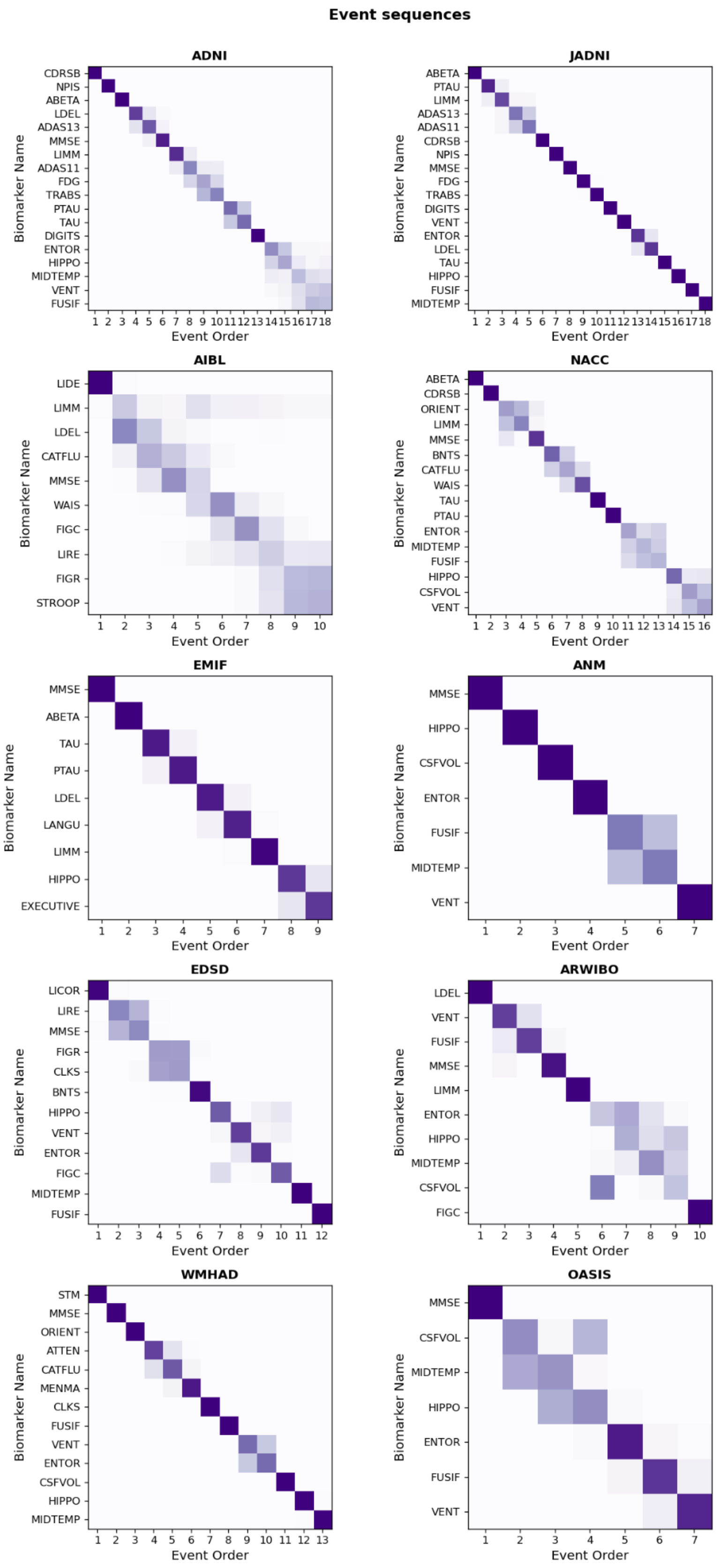


**Figure S2.** The original individual event sequences (independent vertical axes) derived from the ten investigated cohorts.

a)

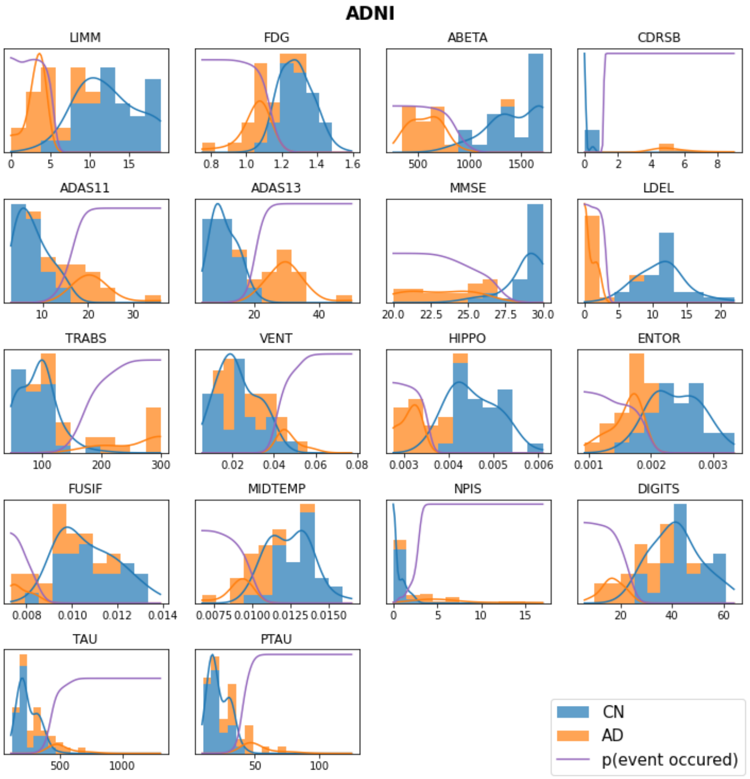
ADNI
LIMM
FDG
ABETA
CDRSB
ADAS11
ADAS13
MMSE
LDEL
TRABS
VENT
HIPPO
ENTOR
FUSIF
MIDTEMP
NPIS
DIGITS
TAU
PTAU
CN
AD
p(event occured)

b)

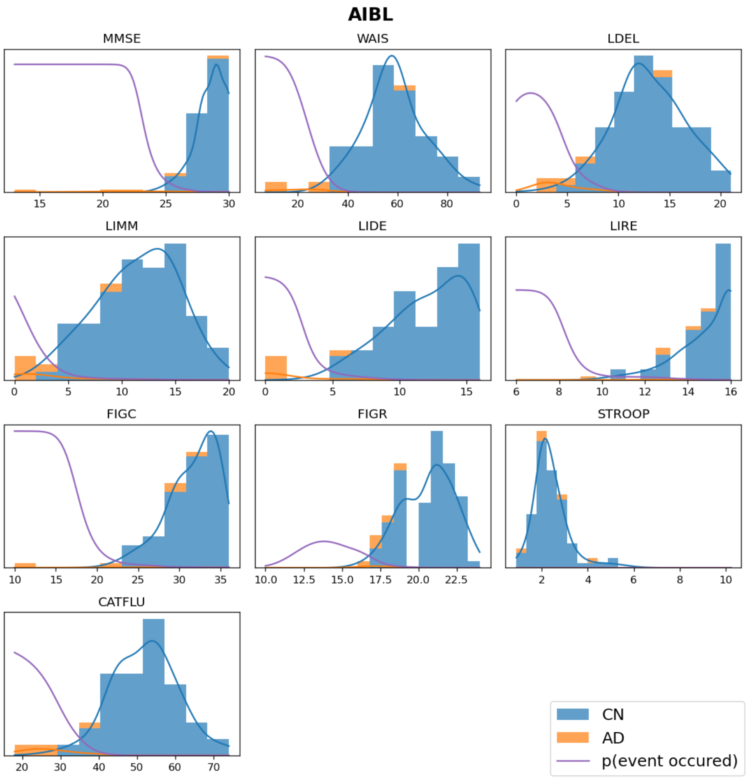
AIBL
MMSE
WAIS
LDEL
LIMM
LIDE
LIRE
FIGC
FIGR
STROOP
CATFLU
CN
AD
p(event occured)

**c)**

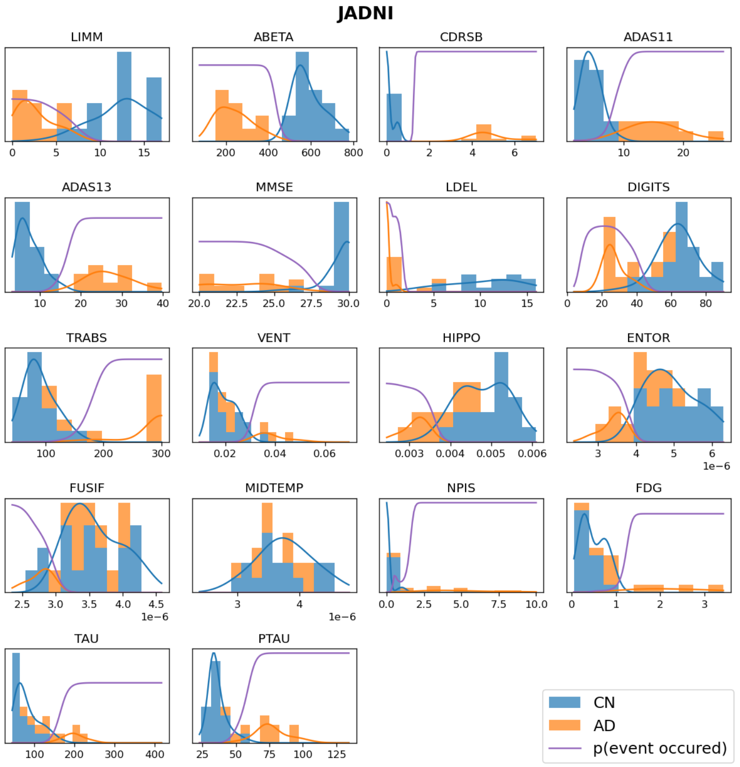
JADNI
LIMM
ABETA
CDRSB
ADAS11
ADAS13
MMSE
LDEL
DIGITS
TRABS
VENT
HIPPO
ENTOR
FUSIF
MIDTEMP
NPIS
FDG
TAU
PTAU
CN
AD
p(event occured)

**d)**

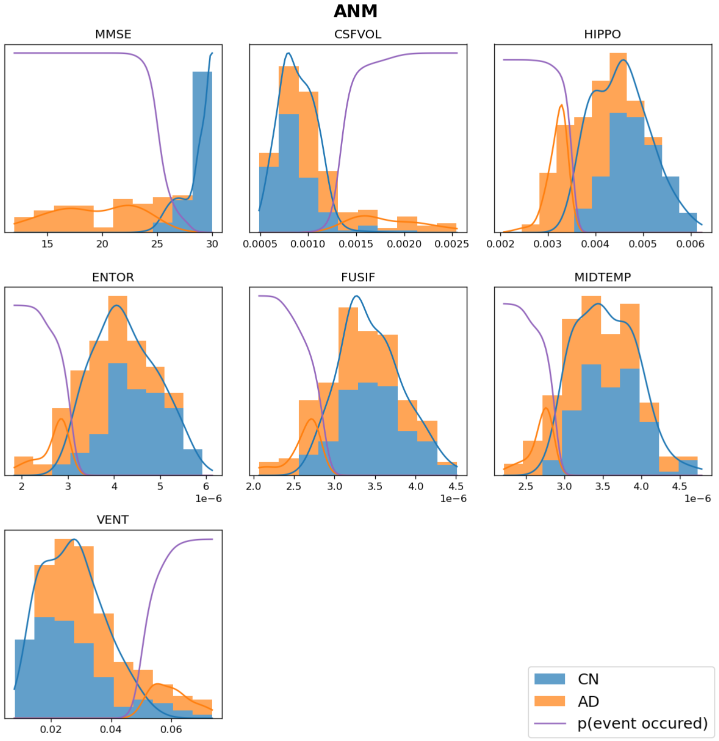
ANM
MMSE
CSFVOL
HIPPO
ENTOR
FUSIF
MIDTEMP
VENT
CN
AD
p(event occured)

e)

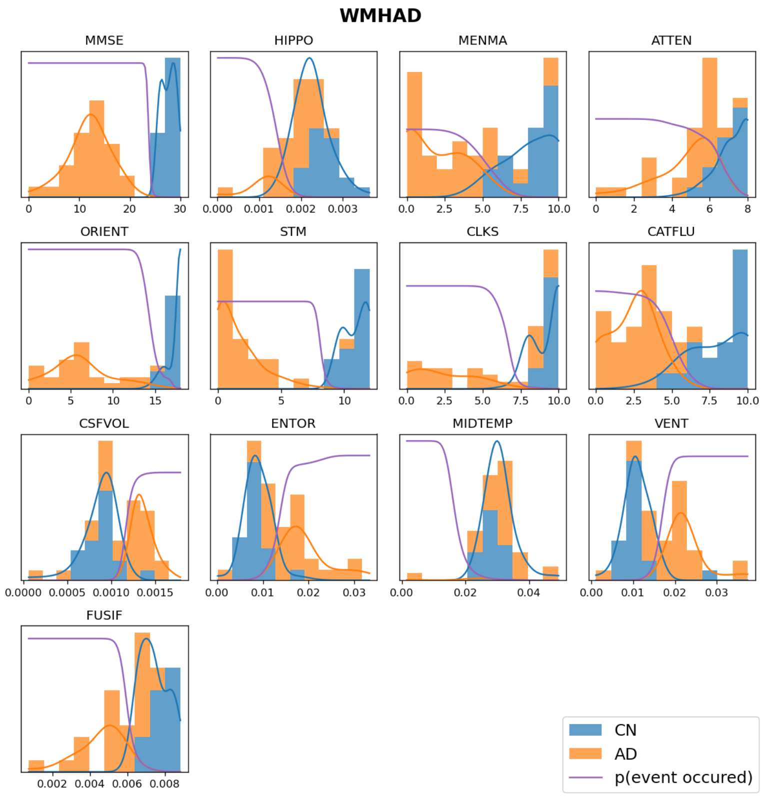
WMHAD
MMSE
HIPPO
MENMA
ATTEN
ORIENT
STM
CLKS
CATFLU
CSFVOL
ENTOR
MIDTEMP
VENT
FUSIF
CN
AD
p(event occured)

f)

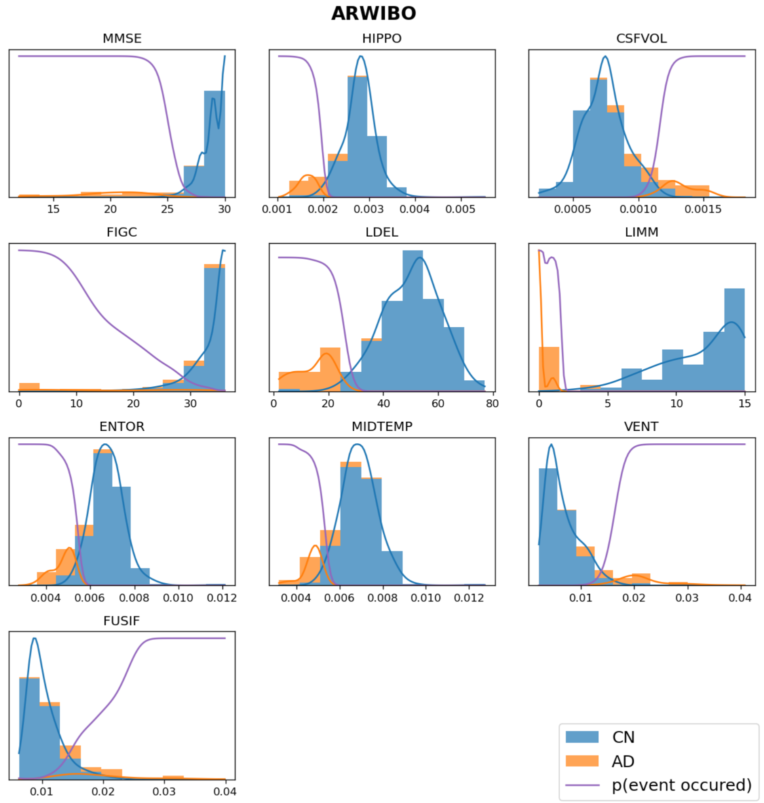
ARWIBO
MMSE
HIPPO
CSFVOL
FIGC
LDEL
LIMM
ENTOR
MIDTEMP
VENT
FUSIF
CN
AD
p(event occured)

g)

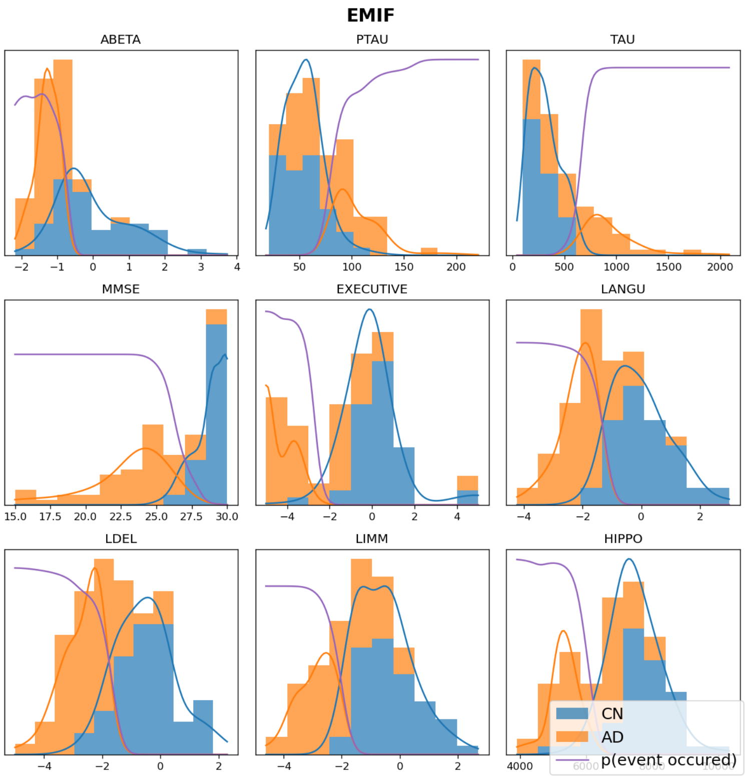
EMIF
ABETA
PTAU
TAU
MMSE
EXECUTIVE
LANGU
LDEL
LIMM
HIPPO
CN
AD
p(event occured)

h)

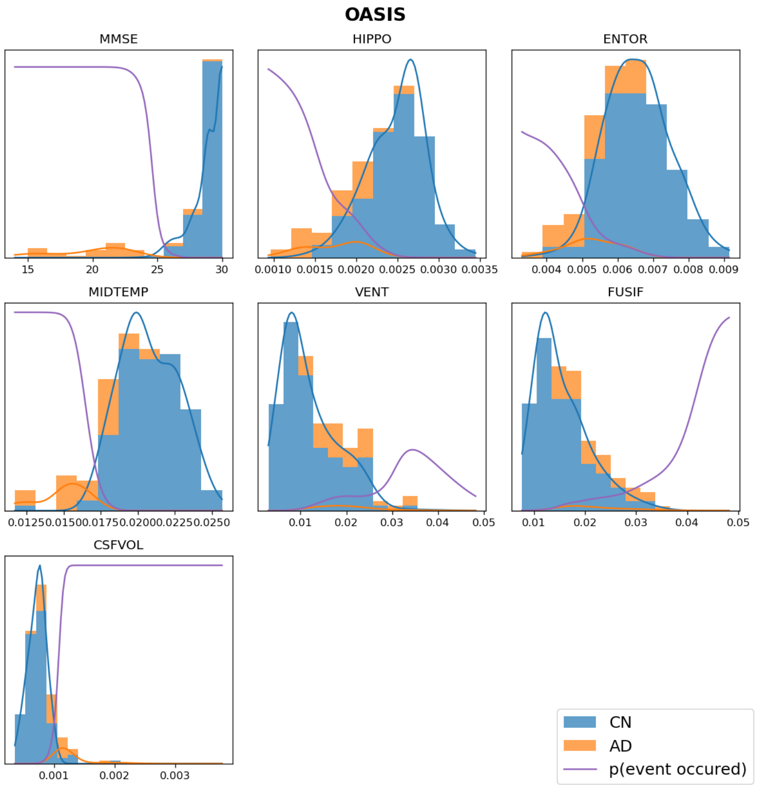
OASIS
MMSE
HIPPO
ENTOR
MIDTEMP
VENT
FUSIF
CSFVOL
CN
AD
p(event occured)

**i)**

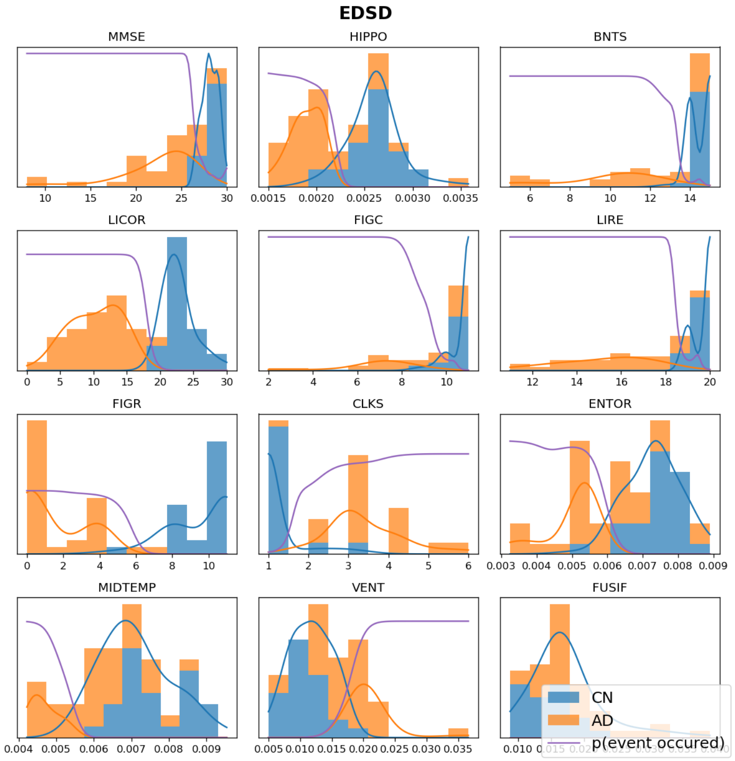


**j)**

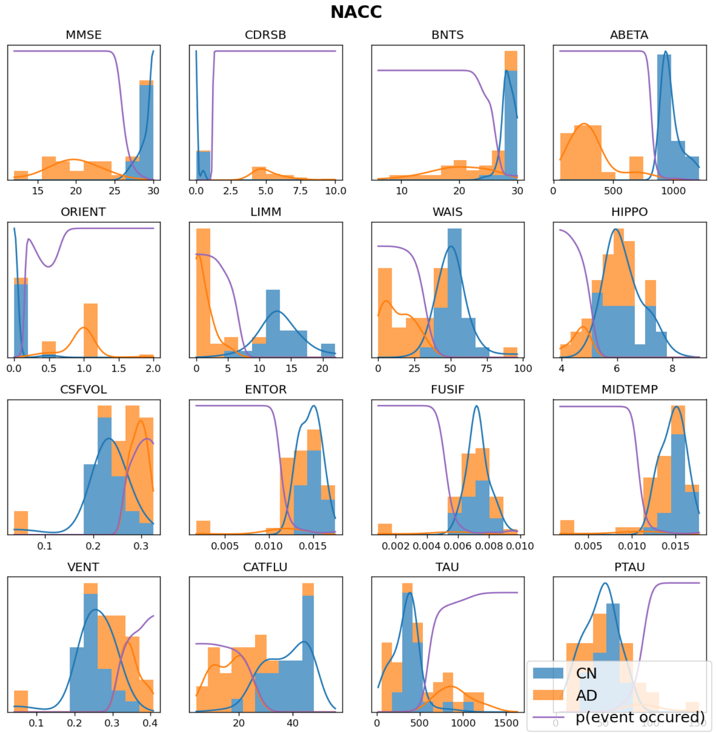


**Figure S3.** The derived mixture models for each cohort (a-j).

### References


1. Fonteijn, H. M., Modat, M., Clarkson, M. J., Barnes, J., Lehmann, M., Hobbs, N. Z., Scahill, R. I., *et al*. An event-based model for disease progression and its application in familial Alzheimer's disease and Huntington's disease. *NeuroImage* 2012, 60(3), 1880–1889. https://doi.org/10.1016/j.neuroimage.2012.01.062
2. Young, A. L., Oxtoby, N. P., Daga, P., Cash, D. M., Fox, N. C., Ourselin, S., Schott, J. M., *et al*. A data-driven model of biomarker changes in sporadic Alzheimer's disease. *Brain* 2014; 2564–2577. https://doi.org/10.1093/brain/awu176
3. Oxtoby, N. P., Young, A. L., Cash, D. M., Benzinger, T., Fagan, A. M., Morris, J. C., Bateman, R. J., *et al*. Data-driven models of dominantly-inherited Alzheimer's disease progression. *Brain* 2018; 141(5), 1529–1544. https://doi.org/10.1093/brain/awy050
4. Firth, N. C., Primativo, S., Brotherhood, E., Young, A. L., Yong, K., Crutch, S. J., *et al*. Sequences of cognitive decline in typical Alzheimer's disease and posterior cortical atrophy estimated using a novel event-based model of disease progression. *Alzheimers dement* 2020, 16(7), 965–973. https://doi.org/10.1002/alz.12083
5. Zhang, W., Zhang, Z., Chao, HC. *et al*. Kernel mixture model for probability density estimation in Bayesian classifiers. *Data Min Knowl Disc* 2018; 32, 675–707. https://doi.org/10.1007/s10618-018-0550-5
6. Scott, D. W. (1979). On optimal and data-based histograms. *Biometrika* 1979, 66(3), 605-610.
7. Li, X., Wang, X., & Xiao, G. A comparative study of rank aggregation methods for partial and top ranked lists in genomic applications. *Briefings in bioinformatics* 2019, 20(1), 178–189. https://doi.org/10.1093/bib/bbx101
8. Lin, S.. Rank aggregation methods. Wiley Interdisciplinary Reviews: Computational Statistics 2010, 2(5), 555-570.
